# FACILITATORS AND BARRIERS TO UPTAKE OF CHILD-FRIENDLY 3 MONTHS OF ISONIAZID AND RIFAMPICIN (3HR) FIXED DRUG COMBINATION (FDC) FOR TUBERCULOSIS PREVENTIVE THERAPY (TPT) IN NIGERIA

**DOI:** 10.1101/2024.09.02.24312961

**Authors:** Rupert Eneogu, Austin Ihesie, Olugbenga Daniel, Ogoamaka Chukwuogo, Debby Nongo, Aderonke Agbaje, Bethrand Odume, Joseph Kuye, Omosalewa Oyelaran, Daniel Egbule, Wayne Van Gemert, Lucy Mupfumi, Cleophas D’auvergne, Obioma Chijioke-Akaniro, Chukwuma Anyaike, Sunday Olarewaju

**Affiliations:** USAID, Nigeria; IHVN, Nigeria; KNCV, Nigeria; John Snow Inc. Nigeria; Stop TB Partnership; USAID Global Health Bureau; Federal Ministry of Health, Nigeria; Department of Community Medicine, Osun State University.

**Keywords:** 3HR (3 months of isoniazid and rifampicin), TPT-TB preventive therapy, Child Contacts, Nigeria

## Abstract

The uptake of TB preventive therapy (TPT) for child and adult contacts of index TB patients in Nigeria has been suboptimal. Nigeria introduced the 3-month isoniazid-rifampicin (3HR) shorter regimen TPT for all eligible contacts through the USAID-funded Stop TB Partnership introducing New Tools Project (iNTP). This study assesses the facilitators and barriers to the uptake of the newly introduced child-friendly 3HR TPT for child contacts of TB patients in Nigeria.

This was a cross-sectional descriptive study using mixed methods. In-depth interviews were conducted among 36 purposely selected Healthcare Workers (HCWs) and 36 caregivers of child contacts. Records of TPT-eligible child contacts (0–14 years) from April to September 2022 were retrospectively extracted. Study data were analyzed using appropriate statistical tools for qualitative and quantitative techniques.

There were 7,906 child contacts identified, of which 7,902 (99.9%) contacts were screened for TB, 1,704 (21.5%) were presumptive, 264 (15.5%) diagnosed with TB, and 6,994 were eligible for TB Preventive Therapy. Additionally, out of 6,994 eligible child contacts, 3984 (57%) were initiated on TPT with 2,982 (74,8%) enrolled on 3HR and 1,002 (25%) on 6H. Of the clients placed on 3HR, 2,499 (85%) completed treatment, 309 (10.5%) lost to follow-up, 27(0.92%) developed Tuberculosis while 48 (1.6%) interrupted treatment.

The key drivers of 3HR TPT uptake among child contacts were TPT-related health education and counseling. Access barriers to 3HR TPT elicited included stigma, poverty, transportation cost, stock out, ineffective monitoring and management of side effects issues of 3HR TPT, subpar Government funding and commitment to addressing TPT implementation challenges and inadequate knowledge among HCWs and caregivers.

In conclusion, 3HR-TPT uptake among child contacts of index TB patients was high. Enhanced provider training and intensive community health education should be sustained while identified individual, structural, and institutional barriers should be addressed to improve implementation.

## Introduction

Nigeria is the leading-burden nation in Africa and the sixth-highest-burden nation for tuberculosis. (1) Particularly among those with HIV, tuberculosis is a prominent cause of death. In 2022, 1.3 million deaths were reported compared to 1.6 million in 2021. (1) As of 2020, only 10% of the expected total number of TB cases in children was notified among children 0–14 years old. (2)

TB preventive therapy (TPT) was introduced by the World Health (3) Organization (WHO) to ensure that those who are at risk of getting TB are “spared from the morbidity and mortality of the disease” (3). Despite WHO recommendations for implementing and adopting TPT in preventing active tuberculosis among high-risk group contacts of TB patients, its acceptance over the years has been suboptimal (4). Understanding the main barriers to TPT among eligible contacts and developing appropriate interventions is necessary within the Nigeria context in meeting specific targets of eligible household contacts on TB preventive treatment as outlined by the United Nations High Level Meeting on Tuberculosis (1).

Common barriers encountered among HCWs handling PLHIVs who do need testing for Latent TB infection before commencement of TPT include limited awareness and inadequate knowledge of the benefits of TPT or even misconceptions about its usage, resulting in missed opportunities to identify eligible individuals and provide them with the appropriate preventive treatment.(5,6)

However, for non-PLHIV eligible household contacts, the situation is compounded by little or limited access to a tuberculin skin test (TST) or interferon-gamma release assay confirm latent TB infection before commencement of TPT. In such instances, HCWs conduct symptomatic screening and laboratory test confirmation of active TB among presumptive TB patients identified, while those screened negative are counseled and enrolled on a TPT regimen.

Other additional challenges from earlier research among non-PLHIV under five contacts besides low awareness were distance to medical services, particularly in underserved or rural areas (5), worries about possible harmful consequences, and the emergence of medication resistance (7).

On the part of caregivers, fear of stigma and discrimination may prevent individuals from seeking preventive therapy, and difficulty in maintaining treatment adherence due to prolonged duration may contribute to poor uptake of Isoniazid TPT (8).

Kenya, Nigeria, and South Africa reported a traditional TPT treatment consisting of 6 to 9 months of daily isoniazid with suboptimal outcomes; however the World Health Organization has since recommended newer medication regimens with shorter durations and better formulations. (4, 9–11)

It would be interesting to see if these factors also contribute to the uptake and acceptance of the 3HR TPT regimen in Nigeria, coming on the heels of the long-standing implementation of the IPT-based regimen. Educational status in a study in Kenya has been shown to influence the uptake of TPT in Kenya, with children of caregivers who had attained education up to the secondary school level 70% less likely to receive IPT than children of caregivers with primary or no educational status (9). The newer, shorter regimen of 3HR TPT has been demonstrated to be safe and well-tolerated in children (12). With its availability in Nigeria, this TPT is expected to be well accepted with improved initiation and completion rates for children (12). Prior research conducted in Ethiopia found that the IPT-based TPT regimen had low completion rates in program-based settings, with initiation rates ranging from 21 to 58% and completion rates of 13% respectively (7).

In addressing these barriers associated with old recommendations of Isoniazid regimen given for six months among under five contacts, the Nigerian Tuberculosis Program received donations of 3HR, including pediatric-friendly formulations (50/75mg dispersible tablets) in late 2021 through the USAID/Stop TB Partnership introducing New Tools Project (iNTP). In early 2022, 3HR pediatric-friendly formulations (50/75mg dispersible tablets) along with adult formulations were distributed to 18 states supported by USAID. Healthcare workers were trained on the use and administration among eligible contacts to improve TPT uptake and reduce the tuberculosis burden. Considering 3HR medications for TPT are a new introduction to the program, this study seeks to assess the facilitators and barriers to the uptake of the newly introduced child-friendly 3HR TPT in Nigeria based on perceptions of HCWs and caregivers. This is expected to influence programming policies and guidelines to improve coverage of TPT for eligible persons.

## METHODOLOGY

### Study Setting

The study was conducted in public and private facilities within the 18 States in Nigeria, supported by the USAID-funded TB-LON project implemented by KNCV Nigeria and IHVN Nigeria. Child contacts (0–14 years old) of index tuberculosis cases living in similar households were traced and screened for TB using a symptom-based approach. Presumptive individuals identified undergo laboratory testing to rule out active TB. Individuals who screened negative for active TB using laboratory tests and those who were not presumptive were enrolled on TPT without confirmation of TB infection from testing due to the non-availability of skin testing and gamma interferon assay. The TPT is monitored monthly till the end of treatments. Documentation was done with the TB contact management register located within the facility. The 18 study states were considered priority states for USAID TB intervention projects. They have a combined estimated population of 114,797,441 persons and an average case notification rate of 124 per 100,000 population in 2021 based on the 2016 population data. The 18 states account for 69% of the national case notification.

### Study Design

Cross-sectional mixed quantitative and qualitative exploratory method was used in this study. For the quantitative component, all children (0-14 years) eligible contacts enrolled and initiated on 3HR between April 2022 and September 2022 had their contact tracing management records retrospectively reviewed. The qualitative component involves conducting an in-depth interview with 36 caregivers and 36 HCWs to understand barriers and facilitators to 3HR TPT.

### Quantitative Data

Relevant data using cascade relating to the uptake of 3HR-based TPT in children and the study outcome was extracted between April 3^rd^, 2023 and April 14^th^, 2023 using the Contact management register between April 2022 – September 2022.

All participants were anonymized before the research team could access them. Personal identifiers like Name, addresses and others were replaced with unique codes as primary key to identify each subject while maintaining confidentiality and living individual participants’ identities private throughout the research process.

### Qualitative Data

This includes consenting HCWs in public and private facilities within 18 TB LON-supported states and consenting caregivers of child contacts of bacteriological diagnosed TB patients on treatment(including TB patients co-infected with HIV) and parents or caregivers of 3HR eligible child contacts of bacteriological positive index TB patients on therapy in the implementing facility, consenting HCWs working in TB DOTS units of 3HR implementing public or private facilities that directly support TB patient management, TB contact investigation administration and management of TPT, HIV negative children between 0-14 years of age who are contacts of bacteriological positive index TB case and child contacts whose data are appropriately recorded and reported in the contact management register of 3HR-TPT implementing facilities. Excluded include TB patients on treatment without household contacts, contacts of adult TB patients who are >14 years of age, contacts of Extra Pulmonary TB patients or contacts of Drug Resistant-TB patients, and HCWs working in other service delivery points in the study facilities, Those who were unwilling to participate in the study as well as those who did not sign informed consent (either in person or by the guardian) linked to the initial statement

### Sample size determination

For the qualitative stage of the study, the sample size was determined by purposive sampling of 4 respondents from all 18 States involved in the study. The respondents include 2 HCWs involved in administering and managing 3HR for TPT and two caregivers with children who were contacts of TB patients and qualified for 3HR TPT. However, the children might not receive 3HR TB preventative treatment.

For the quantitative component of the study, total sampling was used to collate data on the contact investigation cascade (with a focus on the childhood component) for all facilities implementing 3HR TPT in the 18 TB LON project states.

### Sampling technique

All 43,712 medical records of children (0-14 years) who met the inclusion criteria were examined for the study’s quantitative component. Based on the objectives of the research and the characteristics of the population, a non-probability selection technique was employed to choose healthcare providers and caregivers of children, parents, or relatives of children who are contacts of index TB cases on treatment and are eligible for TPT using 3HR contact for the qualitative component’s purposive sampling.

The qualitative phase entails using pre-tested, in-depth interviews that contain information on demographic variables, opinions, and views regarding facilitators and barriers to TPT uptake, personal perspectives on the medication, TPT uptake, and completion rates.

### Data Collection

A data extraction tool for the quantitative aspect was used to extract information using cascade indicators from contact tracing screening activities to focus on the subset of children, identifying eligible child contacts, TPT enrollment, and completion regarding uptake and completion rate among eligible pediatrics contacts of TB index patient between 0-14 years of age between April 1 – September 30, 2022, from the facility TB contact management register.

Similarly, an interview guide was developed using a previous study (2) and pre-tested on 2 HCWs and two caregivers in Ekiti state, modified for later use in data collection.

Eighteen selected data enumerators were trained under the supervision of the research coordinator to conduct interviews with caregivers of selected child contacts and HCWs who provide them TPT. Discussions and interviews were audio-recorded, transcribed, translated into English as needed, and reviewed for accuracy and completeness by bilingual research staff before analysis.

### Data Management and Statistical Analysis

After checking for completeness and accuracy cleaning, the quantitative data were entered and analyzed using SPSS for Windows version 25 statistical software. Descriptive statistics (frequencies and percentages) were done using tables.

Qualitative data verification for accuracy and completeness was done through reading and re-reading by at least two investigators to ensure all recorded information and variations were identified. Audio-recorded data from the in-depth interviews was transcribed verbatim and translated into English. After transcription, codes were developed by two investigators based on the original terms used and matched. The transcripts and notes were analyzed thematically by categorizing them in line with the specific objectives (providers, patients, and resource requirement-related barriers to implementing TPT). The research team presented, discussed, and checked the codes. Tentative categories and subcategories were created from the clustered codes, and subsequently, main themes emerged based on the patterns and relationships between the categories.

### Ethical Consideration

Ethical approval for this study was sought from the National Health Research Ethics Committee of Nigeria (NHREC). In addition, written consent was obtained from the Parent or caregiver after explaining the purpose of the study before the key in-depth interview.

## Result

**Figure 1:**
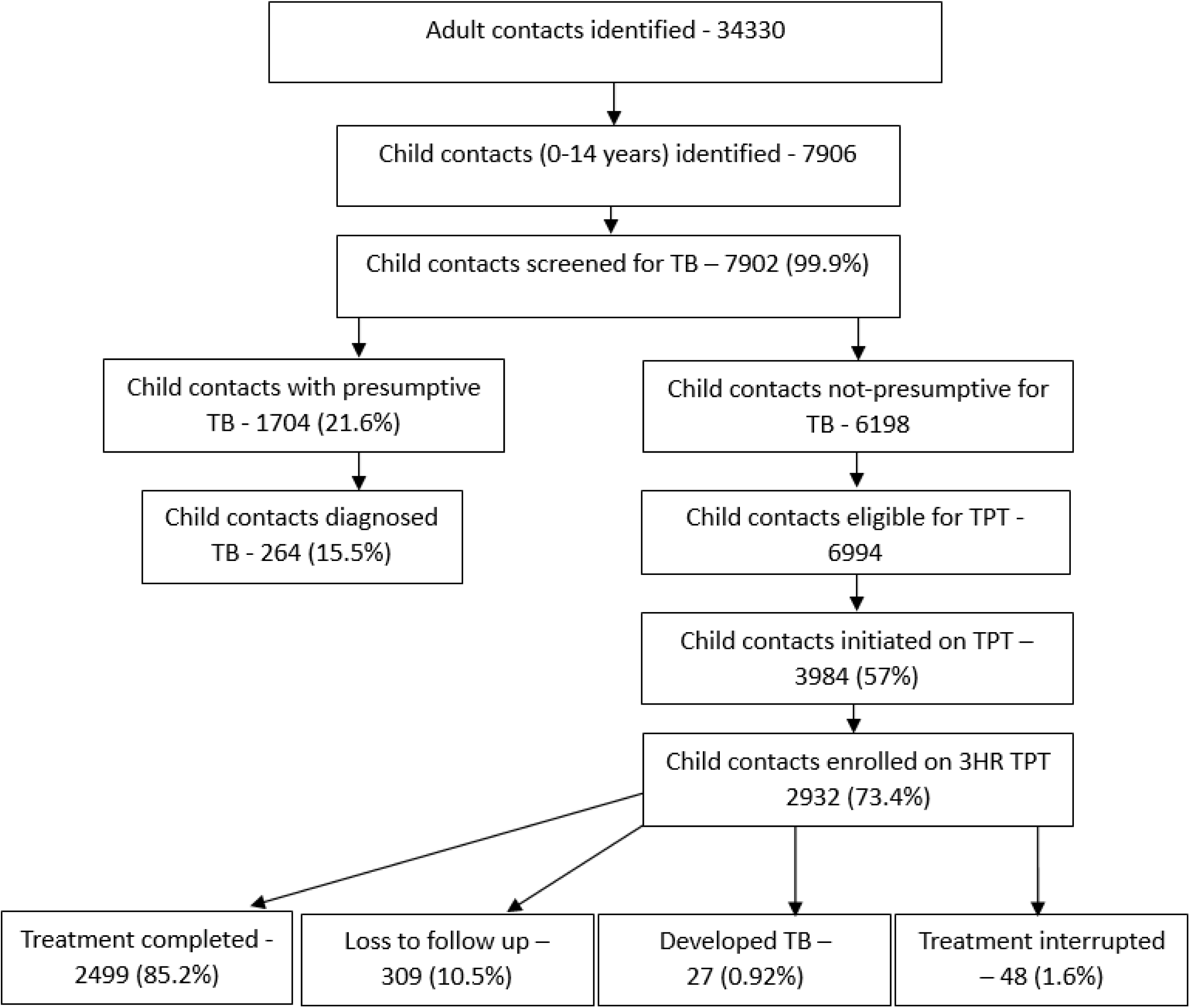
Contact tracing and 3HR cascade among child contacts (0-14 years) April-September 2022, Nigeria.

**Table 1:** Background characteristics of healthcare workers interviewed.

| Variable | Frequency | Percentage |
| --- | --- | --- |
| **Age group** |  |  |
| 20-30 years | 6 | 16.7 |
| 31-40 years | 12 | 33.3 |
| 41- 50 years | 13 | 36.1 |
| >50 years | 5 | 13.9 |
| **Gender** |  |  |
| Male | 9 | 25.0 |
| Female | 27 | 75.0 |
| **Level of education** |  |  |
| Secondary | 2 | 5.6 |
| Tertiary | 34 | 94.4 |
| **Position at the health center** |  |  |
| Nurse | 4 | 11.1 |
| Community Health worker | 7 | 19.4 |
| Others | 18 | 50.0 |
| TBLS officer | 2 | 5.6 |
| DOT officer | 5 | 13.9 |
| **Type of facility** |  |  |
| Public-Tertiary health facility | 9 | 25.0 |
| Public-secondary health facility | 11 | 30.6 |
| Public-primary health facility | 14 | 38.9 |
| Private hospital | 2 | 5.6 |
| **Duration of working experience** |  |  |
| <5 years | 18 | 50.0 |
| > 5 years | 18 | 50.0 |

Out of 36 HCWs interviewed, the majority, i.e., 12 (33.3%), are within 31-40 years. Female 27 (75%), tertiary education (94.4%), working in Public secondary facilities11(38.9%) with working experience of more than five years 18 (50%).

Most participants were younger adults (61.1%) with a mean age of 38.4 years. Similarly, the majority were females (80.6%), had secondary level education (69.5%), self-employed (72.3%), lived more than 1km from the health facility (77.8%), parents (69.5%), and on TB treatment (69.5%). It is worth noting that all (100%) participants had their children on TPT, with 3HR TPT being the most used regimen (72.3%), see Table 2.

**Table 2:**
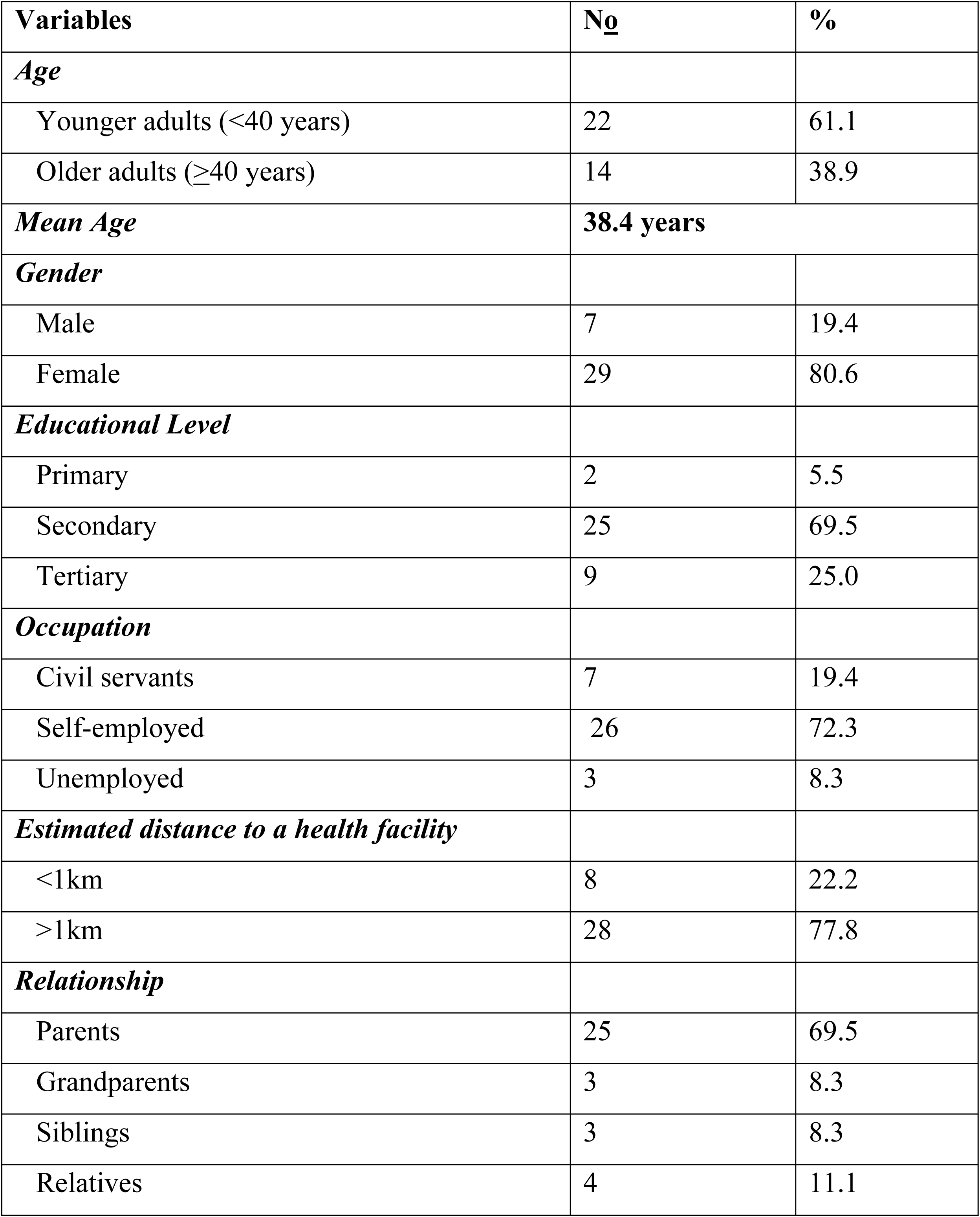
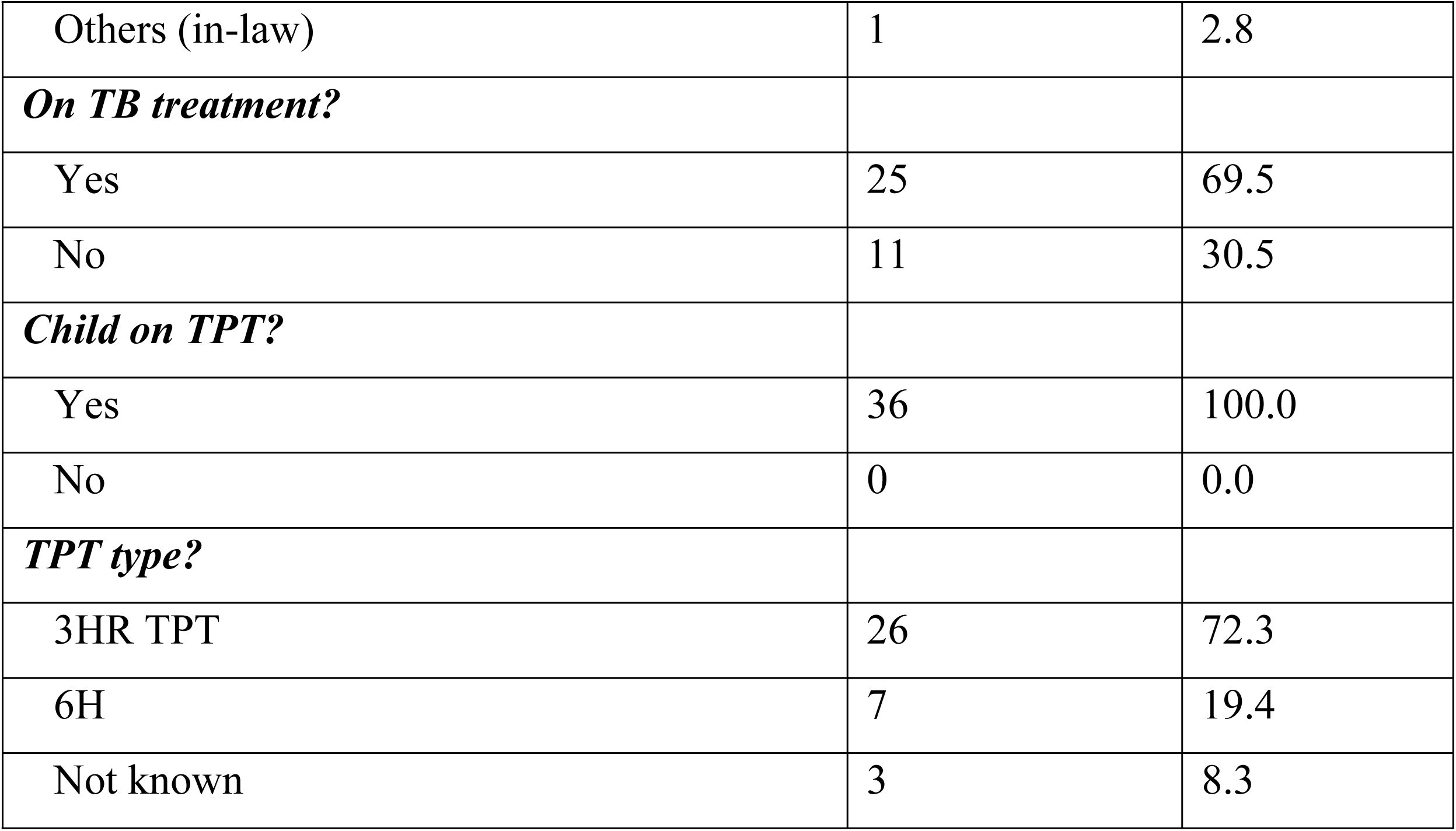
Background characteristics of Caregivers interviewed(N=36)

### Facilitators and Barriers to child-friendly 3HR TPT Enrollment and Possible Solutions

#### Reported Barriers and facilitators to 3HR TPT enrollment with possible solutions among HCWs

Four major themes emerged from the in-depth interviews among HCWs and children’s caregivers as factors perceived as facilitators and barriers to the uptake of child-friendly 3HR TPT among child contacts.

##### 1. Restricted Access to 3 HR TPT

3HR child-friendly TPT is provided freely in public and private health settings; both HCWs and caregivers reported the cost of transportation as part of the structural barrier to accessing TPT for children. Clients are required to visit facilities thrice every month for evaluation by the HCW and monthly refills.

> *“Means of transportation to the facility is a big barrier because it is more of the complaint we receive. DELTA (HCW)*
>
> *“Lagos is busy, and many people complain of being busy and also complaining of traveling, others complain about distance and cost of transportation”* LAGOS (HCW)
>
> *At times, the major challenge we face is transportation fare to come to the facility because we are coming from the village” (**CG1_Katsina, Bauchi, Akwa Ibom, Osun, Rivers, Plateau; CG2_Delta, Bauchi, Anambra) caregiver.***
>
> *“The challenge is coming to the hospital to collect the medication; the cost is the only concern, if we can get support for transport” (**CG2_Osun**) **caregiver.***

Beyond the cost of transportation, other factors hindering initiation and completion of 3HR TPT among children range from negative cultural belief, lack of encouragement from family members, poverty, stigmatization, and discrimination were also reported -

> *“One of the factors that may hinder is a parent not being a biological parent to the child (receiving 3HR) or a nonchalant attitude of the family. ANAMBRA (HCW)*
>
> *“Some don’t use to encourage their family member to come for treatment despite the conditions of the index (case)” CROSS RIVER (HCW)*
>
> *“My family were scared of us, and people ran away from us (child receiving 3HR and Caregiver) thinking they would also contact it” **(CG1_Rivers; CG2_Bauchi) caregiver***

However, it might be possible to lower these costs for some patients and enhance patient enrollment with 3HR TPT services by employing more varied strategies such as the provision of transportation stipends to patients during clinic appointments, reduced treatment appointments, use of home-based drug distribution instead of clinic approach,

> *“I will plead that stipends should be given to index patients and also their caregivers as well.”* LAGOS (HCW)
>
> *“Little stipend to cover transportation for health care; we need the drugs to be always available. Making the drugs in a syrup form”* BENUE (HCW)

Also, clinic-based 3HR TPT service delivery can be refined by supporting HCWs with telephone calls and appropriate, culturally sensitive Information Education and Communication (IEC) materials to create awareness among patients and caregivers during initial and follow-up clinic visitations.

> *“What I think if they are given motivation like recharge card and transportation to the health care workers and patients (Index case).”* DELTA (HCW)
>
> *Creation of awareness” (**CG1_Anambra**) caregiver*
>
> *“The advertisement should continue just like I saw it on the phone; it should not stop so it can help others too” (**CG1_Rivers**). caregiver*

##### 2. 3HR TPT Drug-related factors

Three critical drug-related factors: stock out of 3HR TPT drugs, worry about possible adverse drug reactions, and **3**HR TPT special requirements influenced child-friendly 3HR TPT uptake.

###### a) Stock out of 3HR TPT Drugs

HCWs believed 3HR IPT initiation and completion is linked to constant supply within supported facilities. Still, some interviewed reported stockouts and shortages of 3HR TPT drugs, creating difficulty in administering TPT to children.

> *“Unavailability of the 3HR “OGUN (HCW)*

To avoid stock-outs and promote an uninterrupted uptake of TPT, HCWs believed it is necessary to maintain a consistent and adequate supply of medications through training and job aids on drug logistic management.

> *“Training and provision of the manual for easy understanding so that each DOT Officer will read and understand each case treatment.”* KADUNA (HCW)
>
> *“Let us have enough drugs in order not to get stockout; it will help a lot in the uptake”* KATSINA (HCW)

###### b) 3HR TPT side effects

Even though a child-friendly 3HR regimen is usually well tolerated, some children experience unfavorable drug reactions (side effects), which can hinder uptake, according to HCWs and caregivers interviewed.

> *“Its’ effect is minimal, little abdominal discomfort for a few periods” (**CG2_Osun**) Caregivers must know during counseling sessions from HCWs about adverse effects and their management measures during initial start-up and follow-up monthly visitations.*
>
> *“It still goes back to good counseling; what are the benefits and the reason for that? You need to do a groundwork for a better response.”* KADUNA (HCW)

###### c). 3HR TPT special requirement for children

Children frequently have special requirements and challenges, according to caregivers interviewed, such as increased appetite after drug use and reluctance to take the medication/ *“They (the children) only complain of hunger after taking the drug” (**CG2_Rivers; CG1_Lagos, and Ogun**)*

###### c) 3HR TPT Monitoring measures

After initiating 3HR TPT, HCWs must monitor adherence, monitor side effects, and ensure completion. HCWs interviewed reported using personal money to send text message reminders and conducting personal home visits.

> *“Yes, they are taking it; we used to monitor from time to time, calling their parents (caregivers) to find out from them.”* BENUE (HCW)
>
> *We used to monitor them, visit them at home, and also give them drugs at home.* KADUNA (HCW)
>
> *I used to call the clients (caregivers), and I used my money to call them. I asked them about the progress in their health status. AKWA IBOM (HCW)*

##### 3. Existing gap in both human and non-human resources concerning 3HR TPT Policy service delivery

Quality of services delivered within supported facilities is linked to workload per staff and other support provisions by the Government and other stakeholders. In this study, HCWs reported a shortage of manpower and other logistics support required for adequate service provision.

> *“Lack of manpower and labor force to facilitate more work and coverage.”* ANAMBRA (HCW)
>
> *“Lack of sufficient staff in the facility to aid in counseling and support in the TB units, provision of some stipends to the workers of TB units.”* OSUN (HCW)
>
> *“Lack of encouragement from the government”* OYO (HCW)

##### 4. 3HR TPT-related health education and counseling

Inadequate knowledge about the advantages and safety of 3HR TB TPT for children was seen as a barrier to the successful uptake and completion of child-friendly 3HR TPT by HCWs and caregivers.

> *“Some of the caregivers don’t know about the drugs”* ANAMBRA (HCW)
>
> *“Lack of proper awareness to the parent (caregiver) can lead to such.”* KATSINA (HCW)

To address ignorance, HCWs acknowledged the importance of training to enhance their ability to administer treatment effectively and provide proper counseling sessions during initiation and follow-up visitation.

> *“Relationship establishment with the patients that will encourage him to take the drugs”* ANAMBRA (HCW)
>
> *“Method of counseling should be simple and acceptable and also explain the importance of the drugs to them” KANO (HCW).*
>
> *“Training of health workers is very key to placing people on treatment well and counsel them effectively.”* BAUCHI (HCW)

### Framework for reported barriers and possible solutions to uptake of child-friendly 3HR TPT in Nigeria

### Factors enhancing successful child-friendly 3HR TB Preventive Treatment among HCWs and caregivers in Nigeria

A successfully delivered child-friendly 3HR TPT requires adequate knowledge through enhanced training on the part of HCWs coupled with community TPT-related education and counseling sessions among caregivers to dispel myths and misconceptions -

> *“It (3HR TPT) is very effective in children, we appreciate the drugs as it assists the children, it is safe because it is well packed and organized.”* NASSARAWA (HCW)
>
> *It (3HR TPT) will help to protect the children against TB.”* LAGOS (HCW)

Additionally, HCWs and Caregivers reported the detrimental effects of TB, the value of the protection that child-friendly 3HR TPT offers to children exposed to TB, and the significance of 3HR TPT in the fight against the disease during education and counseling sessions with index TB cases

> *“I used to educate them on the need for them to take the drugs and not miss it.”* OSUN (HCW)
>
> *“I counsel them on the use of the drugs and the importance of taking the medication for prevention and not to default”* KANO (HCW)
>
> *“It (TB) can be prevented by going to the clinic and getting your TB TPT and the use of face mask” **(CG1_Delta, Imo, Rivers, and Plateau; CG2_Akwa Ibom) Caregiver***
>
> *“I was told about the importance of the medication and why we should start the treatment immediately” (**CG2_Rivers; CG1_Lagos**)*
>
> *“It (3HR TPT) is very reliable and can be trusted. It is given to children to prevent them from being infected with TB because their body is still fragile, they are prone to diseases” (**CG1_Crossriver, Bauchi, and Plateau; CG2_ Bauchi, and Anambra**) Caregiver*

Moreover, the ease of administration with short duration and sweetness were other factors reported by caregivers contributing to the successful uptake of child-friendly 3HR TPT

> *(3HR Child-friendly TPT) was not bitter is just like sweet” (CG2_Ogun, and Delta)*
>
> *(3HR Child-friendly TPT) is very effective and easy to use” (CG2_Osun, and Bauchi) “And it is a 3-month regimen” (CG1_Crossriver, Benue, Kaduna, and Osun)*

There were no appreciable differences between the health care providers and caregivers about what makes a program successful.

## DISCUSSION

Understanding the perceptions of index patients, caregivers, and HCWs about the recently introduced 3HR TPT regimen will help the National TB control program meet its goals for acceptance and completion goals in line with the Stop TB Strategy.

This study, which we believe to be the first of its kind in Nigeria, used quantitative and qualitative techniques to explore perceptions that will either facilitate or act as a barrier to the uptake of the new 3HR TPT regimen, particularly among caregivers of child contacts and HCWs in Nigerian hospitals concerning children within 0-14 years old.

Documentation from existing health records between April and September 2022 of the child contacts placed on TPT, 73.4% of them were initiated on 3HR TB preventive therapy. This is higher than 66% reported in Kenya by Ngugi et al. among HIV-positive children (1-9 years) using six months of TPT and 64.3 % in Ethiopia by Tadese et al. among under-five HIV-positive children (4,7).

Similarly, a Nigerian study on the practice of isoniazid preventive treatment in Ebonyi States among adults that are HIV positive had a lower percentage, which was 17% (10).

The variation compared to our study outcome could be due to differences in settings. In addition, the higher rate reported in our study could be attributed to capacity-building activities that HCWs underwent in the form of training, mentorship, and supervision to improve their knowledge of the safety and benefits of 3HR TPT. This helped them to provide better counseling to caregivers during the initial and subsequent monthly visits to dispel myths and misconceptions, as acknowledged from the results of the qualitative survey.

Furthermore, regarding treatment outcomes for the 3HR regimen among child contacts, 85% completed treatment, 10.5% were lost to follow-up, 0.92% developed Tuberculosis, and 1.6% interrupted treatment. Previous studies reported completion rates of 3.7% in South Africa, 20% in India and 67.9% in Ethiopia using six months of isoniazid (13–15).

Based on the information from the in-depth interview, the higher completion rates reported in this study can be attributed to the significantly shorter treatment duration, TPT-related health information learned during counseling sessions by caregivers from HCWs, and possibly due to the pleasant child-friendly formulation of the drug that encouraged adherence and completion. This study discovered that the dedication to enhancing the uptake of child-friendly 3HR TPT among HCWs and caregivers in Nigeria was linked to continuous health education and counseling geared towards convincing caregivers and child contacts to adopt behaviors that are good for their health. Health education is a crucial activity for ensuring adherence. When index TB cases and child caregivers comprehend how 3HR TPT helps prevent TB, they can better make educated judgments and accept the intervention when offered. Furthermore, the need for additional enhanced 3HR TPT training coupled with a simplified National algorithm among HCWs, more 3HR TPT community-based education, and counseling sessions to address myths and misconceptions would help caregivers and child contacts understand the value of protecting overall well-being. Additionally, the ease of administration of child-friendly 3HR TPT with short-duration was another facilitating factor reported by caregivers during the in-depth interview. Similarly, previous studies demonstrated high treatment success using three or 4- month regimens of isoniazid and rifampicin or rifampicin alone. (16–19).

The program’s implementation strategy in our study showcased the effectiveness of employing contact tracing officers for focused Contact Investigation and follow-up. This approach enabled the identification of child contacts linked to bacteriologically confirmed index TB patients. By conducting thorough screenings, presumptive cases were identified and subsequently differentiated into confirmed TB cases requiring treatment and those without symptoms or negative presumptive evaluation results, who were placed on TB preventive therapy.

The in-depth interviews with healthcare workers (HCWs) and caregivers unveiled four key barriers impeding the uptake of 3HR TPT among child TB contacts. These include challenges related to Inadequate health education and counseling specific to 3HR TPT, Limited access to TB services, factors associated with the 3HR TPT drug regimen, and Insufficient Human and Non-Human resources for implementation at the facility level (see Figure 2). Notably, health education and counseling emerged as pivotal elements influencing caregivers to enroll the child contacts in and complete 3HR TPT. This underscores the critical role of educating individuals about the intervention’s effectiveness, benefits, and safety in dispelling myths and misconceptions, thus helping them make educated judgment to readily accept TPT. This aligns with prior studies in Nigeria and Kenya by Chizoba et al., and Nwangi et al., which show education has been consistently linked to improved uptake of TB preventive therapy (9,10).

**Figure 2.**
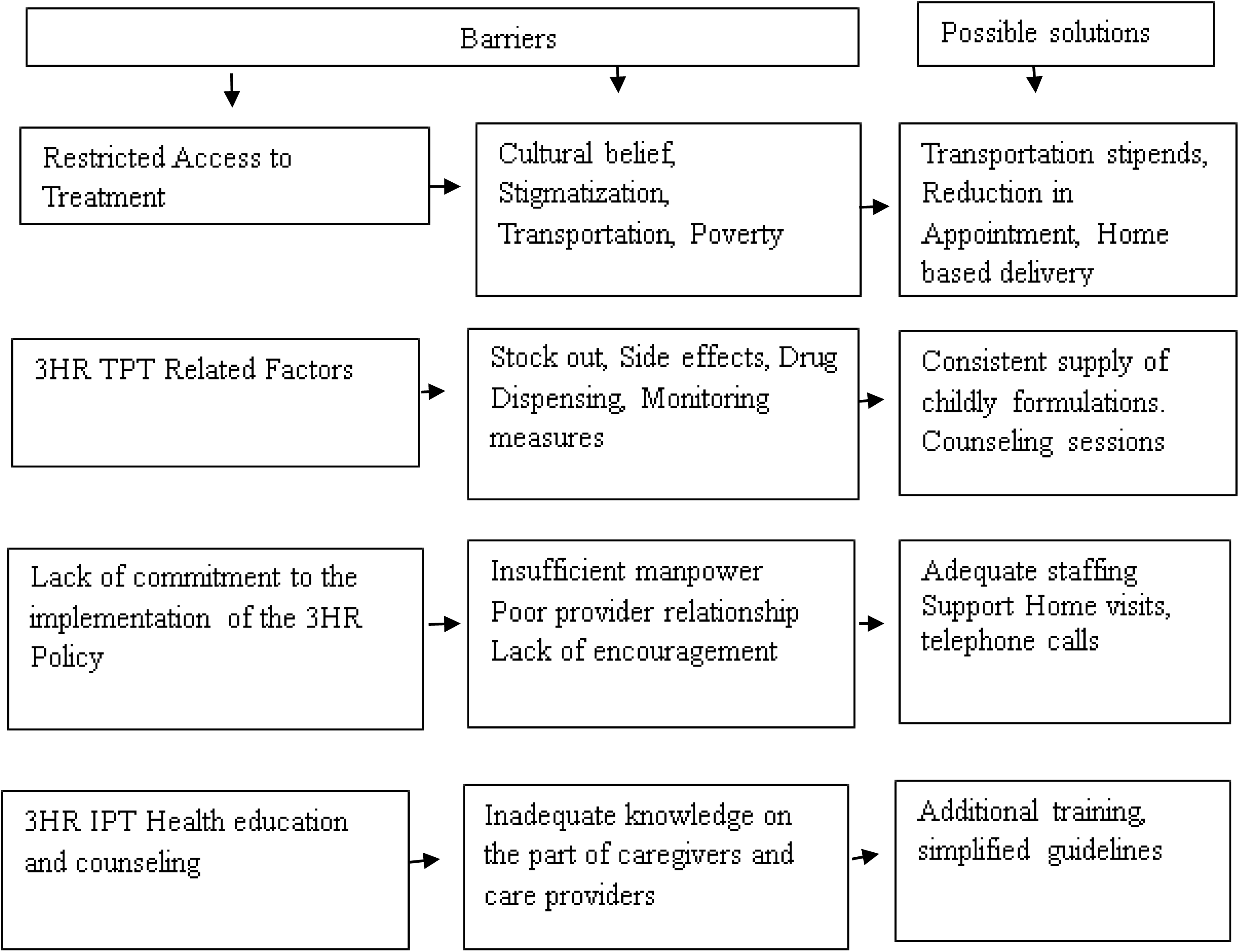
Driver diagram in identifying barriers to uptake of child-friendly 3HR therapy among HCWs and care providers with possible solutions.

Despite the availability of free child-friendly 3HR TPT in both public and private health settings, caregivers and HCWs identified the cost of transportation as a significant structural barrier to access. This is consistent with findings from earlier studies by Assefa et al. in Ethiopia and Rekha et al. in India. ((20,21) which highlights the above amongst other factors responsible for the poor TPT uptake. The recommendations from respondents in our study include providing transportation stipends, reducing treatment appointments, adopting home-based drug initiation (Community Initiation), and facilitating drug dispensing through the nearest health facility. In addition, respondents in our study highlighted stigma and discrimination as one of the challenges affecting treatment. This is similar to reports from Ebonyi, South Eastern Nigeria, which identified stigma and discrimination by family members as hindrances to TB preventive treatment. (10)

This justifies the need to involve patient relatives and community members in promoting acceptance and adherence.

Additionally, potential barriers such as drug stockouts, adverse side effects, and specific needs related to drug dispensing and monitoring within the three-month period were identified. In alignment with prior research findings, these challenges can be mitigated through effective drug supply management and comprehensive patient/caregiver education as guided by healthcare workers’ prescriptions, in alignment with prior research findings. Also, side effects of peripheral neuropathy reported can be reduced by adequate patient/caregiver health education on using Vitamin B6 based on HCWs prescription as suggested by other studies. (22)

Another barrier to successful implementation was the shortage of HCWs required to provide 3HR and the lack of incentives in supporting logistics required for effective child-friendly 3HR TPT. In the spirit of ownership and sustainability, the government should provide stipends to support communications and home visits to HCWs during tracking and identifying TB child contacts of index patients, enrollment, and monitoring of 3HR TPT treatment.

Our quantitative evaluation using the existing TB contact management register has some limitations as a research tool. Some cases with missing variables were removed from the study, leaving only patients with complete and accessible records. Therefore, a particular interest group may have been underrepresented by the study. Also, some candidates who refused treatment despite HCWs’ counseling were excluded. Additionally, participants in the qualitative study were predominantly female.

Nevertheless, the study has several strengths. First, the qualitative aspect thoroughly examined HCWs ranging from nurses and community extension health workers across Primary, Secondary, and Tertiary health facilities and their caregiver perspectives on issues related to facilitators and barriers to 3HR TPT treatment. The selection was done in 18 States across the country’s six geopolitical zones. Qualitative data obtained had multiple layers of quality assurance monitoring to guarantee the reliability of the qualitative interviews.

### Conclusion and Recommendation

3HR TPT uptake and completion is high among childhood contacts of index bacteriologically confirmed TB cases in Nigeria due to the interest and dedication of HCWs and caregivers. The critical facilitators of 3HR TPT uptake were TPT-related health education and counseling, ease of administration, shorter duration, and sweetness of child-friendly TPT drugs. Barriers identified that restricted access to treatment included stigmatization, poverty, and transportation costs. Additionally, other factors affecting the quality of services, such as stock out, side effects of 3HR TPT, existing gaps in both human and non-human resources required in providing effective follow-up of child contacts, as well as inadequate knowledge on the part of HCWs and caregivers should be addressed.

## Data Availability

The corresponding author may provide the data supporting the findings of the research upon reasonable request but, the data are not publicly available. Dr. Sunday Olarewaju can be contacted at with requests for access to data. Justification for Selecting Data Availability Upon Request for Our Manuscript In alignment with ethical research practices and a commitment to transparency, I am making the data from this study available upon request. This decision is rooted in several key considerations: 1. Ethical Responsibility: Protecting participants’ privacy and confidentiality is paramount. By providing data upon request, I ensure that sensitive information is shared responsibly in accordance with ethical guidelines and participant consent agreements. 2. Transparency and Reproducibility: Sharing data fosters transparency and allows other researchers to validate findings, contributing to the integrity of the scientific process. However, I aim to balance this with the necessity of protecting participants’ identities and sensitive information. 3. Data Integrity: By controlling access to the data, I can ensure that it is used appropriately and responsibly, enabling me to maintain oversight on how the data is utilized in subsequent research. 4. Institutional Compliance: This approach aligns with my institution's policies regarding data sharing and participants’ confidentiality, ensuring that I adhere to established ethical standards. I believe this approach supports the advancement of knowledge while upholding the highest ethical standards in research. Thank you for considering this justification

## Acknowledgement

The authors acknowledge the support of the National Tuberculosis and Leprosy Control program, State TB control Managers across the 18 USAID TB-LON supported states in Nigeria, Tuberculosis Local Government supervisors, Health care workers in the selected sites and study participants for their contribution to the success of this study. We also appreciate the technical support and input from the Stop TB Partnership introducing New Tools Project (iNTP) team and USAID Washington TB Division.

## Funding

The study was funded by the United States Agency for International Development (USAID) through the Stop TB Partnership introducing New Tools Project (iNTP).

## Author’s contribution

Conceptualization and design; Rupert Eneogu, Austin Ihesie, Olugbenga Daniel, Ogoamaka Chukwuogo, and Joseph Kuye. literature review; Austin Ihesie, Joseph Kuye, and Daniel Egbule. Study Protocol; Rupert Eneogu, Austin Ihesie, Olugbenga Daniel, Ogoamaka Chukwuogo, Debby Nongo, Aderonke Agbaje, Bethrand Odume, Joseph Kuye, Daniel Egbule, Wayne Van Germert, and Lucy Mupfumi. Data collection; Rupert Eneogu, Austin Ihesie, Olugbenga Daniel, Ogoamaka Chukwuogo, and Sunday Olarewaju Research coordination; Rupert Eneogu, Austin Ihesie, Olugbenga Daniel, Ogoamaka Chukwuogo, Debby Nongo, Aderonke Agbaje, Bethrand Odume, Omosalewa Oyelaran, and Sunday Olarewaju Statistical analysis; Sunday Olarewaju, Writing first draft; Rupert Eneogu, Austin Ihesie, Olugbenga Daniel, Ogoamaka Chukwuogo, Debby Nongo, and Sunday Olarewaju. Revision of draft/approval; all authors.

## Notes

### Competing Interest Statement

The authors have declared no competing interest.

### Clinical Protocols

https://docs.google.com/document/d/1x5dexusYcyeXFToGDEZu8S_UBdKcm9tp/edit?usp=sharing&ouid=115246320401680973472&rtpof=true&sd=true

### Author Declarations

IRB NAME: National Health Research Ethics Committee of Nigeria NHREC Approval Number: NHREC/01/01/2007-29/04/2023 Date: 29th of April, 2023

